# ICU Readmission Prediction for Intracerebral Hemorrhage Patients using MIMIC III and MIMIC IV Databases

**DOI:** 10.1101/2025.01.01.25319859

**Authors:** Hexin Li, Rachel Monger, Elham Pishgar, Maryam Pishgar

## Abstract

**Background:** Intracerebral hemorrhage (ICH) is a critical form of stroke resulting from bleeding within the brain, with a mortality rate of 40-50% within a few days and significant risk of long-term disability. Despite the high incidence of ICU readmissions among ICH patients, the specific factors contributing to these readmissions remain unclear. This study utilizes MIMIC-III and MIMIC-IV databases to develop machine learning models that predict ICU readmissions in ICH patients.

**Methods:** Data from 2,144 patients were extracted using ICD-9 and ICD-10 codes. Four machine learning models - AdaBoost, Random Forest, XGBoost, and LightGBM - were implemented. Recursive Feature Elimination with Cross-Validation reduced features from 50 to 18 key predictors. The RandomUnderSampler technique addressed class imbalance by reducing majority class samples to 60% of the minority class, while the Optuna framework with Tree-structured Parzen Estimator optimized model parameters. Performance was primarily evaluated using AUROC, which effectively handles class imbalance and provides threshold-independent assessment, complemented by accuracy, sensitivity, and specificity metrics.

**Results:** The AdaBoost model achieved an AUROC of 0.877 (95% CI: 0.815-0.913) and accuracy of 0.810, improving from the previous best AUROC of 0.736. Sensitivity notably increased from 22.6% to 84.0%, demonstrating substantial improvement in identifying high-risk patients. This enhancement resulted from effective class imbalance handling and AdaBoost’s adaptive weighting mechanism. Through comprehensive SHAP and ablation analyses, we identified oxygen saturation and cardiovascular disease as crucial predictive features for ICU readmission risk assessment, providing new insights into patient monitoring and care management.

**Conclusions:** Our preprocessing methodology and model selection strategy significantly improved high-risk ICH patient identification. Through comprehensive improvements combining advanced hyperparameter optimization, balanced sampling techniques, and dual feature importance analysis, we achieved a 19.2% AUROC improvement while reducing feature dimensionality from 51 to 18. This integrated approach demonstrates the potential of machine learning in enhancing clinical decision-making. The framework provides a promising foundation for developing clinical decision support tools in ICU settings, improving resource allocation and enabling more personalized patient care interventions.

## 1. INTRODUCTION

Intracerebral hemorrhage (ICH), which occurs due to bleeding within the brain, is both a common and highly fatal form of stroke. With a mortality rate ranging from 40% to 50% in the initial days following the stroke, ICH remains a critical condition. While accounting for approximately 10-20% of all stroke cases, its severity and the associated poor outcomes set it apart from other types of strokes [1]. Despite the overall mortality for ICH remaining at 30% to 40% during the early phase, there has been little advancement in improving these outcomes in recent years [3, 4]. ICH patients also typically face worse neurological outcomes and a significantly higher likelihood of long-term disability when compared to those with ischemic stroke [5].

The severe nature of ICH often leads to prolonged hospital stays, which heightens the risk of various complications. According to [6, 7], in the early stages of the condition, hematoma expansion represents a serious concern. As the disease progresses, patients may develop further complications stemming from underlying health conditions or problems that arise during their hospitalization, such as infections or deep vein thromboses. [8]

Managing ICH patients in Intensive Care Units (ICUs) presents substantial challenges, particularly due to the elevated risk of ICU readmission resulting from delayed medical complications. These readmissions not only lead to worse health outcomes but also contribute to higher healthcare costs [2]. While research has extensively focused on the acute phase of ICH, studies specifically exploring the reasons behind ICU readmissions, including post-acute complications and other less-understood factors, remain limited [9, 10]. Moreover, there is no consensus on the ideal timing for transferring ICH patients out of the ICU. Such decisions often depend on the physician’s clinical judgment and the preferences of the patient or their family. Transferring patients prematurely or delaying the process may lead to higher medical costs, misallocation of critical resources, and an increased risk of complications [11].

Most studies have not fully leveraged advanced machine learning algorithms or effectively addressed the issue of class imbalance in ICH patient care, leaving a significant gap in the literature. The clinical context and feature selection play key roles in evaluating both short-term and long-term factors [12, 21, 16]. while machine learning techniques, in combination with existing healthcare data frameworks, can contribute to more effective risk factor assessment [17, 18]. To address these challenges, our study proposes an integrated framework combining RandomUnderSampler for class imbalance and RFECV for comprehensive feature selection. The key advantages of this approach include: (1) applying rigorous data preprocessing techniques including mean imputation and label encoding; (2) employing a comprehensive feature selection process that combines machine learning techniques with clinical expert insights; (3) effectively addressing class imbalance by reducing majority class samples to 60% of the minority class; (4) implementing advanced algorithms such as AdaBoost, Random Forest, XGBoost, and LightGBM for their superior handling of categorical variables and computational efficiency.

Our study aims to demonstrate the clinical applicability of this balanced prediction framework and evaluate the performance of different machine learning models, particularly the AdaBoost model, in ICU readmission prediction for ICH patients. By carefully evaluating feature selection criteria using data from both MIMIC-III and MIMIC-IV databases, we focus on identifying factors that might influence ICU readmission. Our prediction model complies with the standards of the Transparent Reporting of a Multivariable Prediction Model for Individual Prognosis or Diagnosis (TRIPOD) initiative, guaranteeing thorough and transparent reporting [19, 20].

### 1.1. Statement of Significance

- **Problem:** ICU readmission prediction faces challenges in model interpretability and high false negative rates, limiting clinicians’ ability to effectively identify high-risk patients.
- **What is already known:** Existing machine learning methods for ICU readmission prediction often prioritize accuracy over interpretability, acting as “black boxes” with insufficient sensitivity for clinical application.
- **What this paper adds:** We present an interpretable AdaBoost model with integrated SHAP analysis that achieves improved sensitivity in identifying readmission risks. Our analysis reveals hematological parameters and cardiovascular disease status as key predictors, providing clinicians with actionable insights for risk assessment.

## 2. METHODOLOGY

### 2.1. Data Description

We retrieved past patient data from the Medical Information Mart for Intensive Care III (MIMIC-III, version 1.4) and the Medical Information Mart for Intensive Care IV (MIMIC-IV, version 2.0) databases [23, 22]. Both of these are large, single-center databases from the Beth Israel Deaconess Medical Center between 2001 and 2012. The data within these databases are deidentified and consist of an abundance of data such as laboratory results, vital signs, demographics, medications, and more.

### 2.2. Patient Inclusion Criteria

Using ICD-9 code (431) and ICD-10 codes (I610-I616, I618-I619), we identified patients with Intracerebral Hemorrhage through SQL queries in BigQuery, as shown in Fig1. Only patients with ICH as either primary or secondary diagnosis were considered for this study.

**Figure 1.**
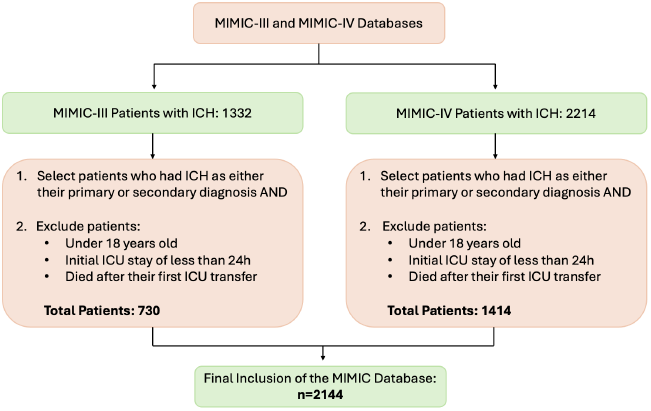
Patient Extraction Criteria

The following exclusion criteria were applied: (1) Age under 18 years; (2) Initial ICU stay less than 24 hours; (3) Death after first ICU transfer; and (4) For patients with multiple ICU admissions, only the first admission was considered.The final cohort comprised 2,144 patients, with 730 from MIMIC-III and 1,414 from MIMIC-IV databases. These patients were combined into a single dataset for subsequent train-test splitting.

### 2.3. Feature Extraction

The feature selection process in our study involved several stages. Initially, through comprehensive literature review [1, 12, 37, 38, 13] and expert insights, we identified 50 potential features encompassing various clinical and demographic aspects of ICH patient care. These initial features included vital signs, laboratory measurements, comorbidities, clinical assessments, and treatment-related factors.

To optimize our feature set, we applied RFECV to identify the most significant predictors. This process helped reduce the dimensionality from 50 to 18 key features by evaluating each feature’s contribution to model performance. The selection threshold was determined through cross-validation, ensuring optimal balance between model complexity and predictive accuracy. The final selected features included demographics (age, gender), vital signs (heart rate, blood pressure, respiratory rate, oxygen saturation, Glasgow Coma Scale), laboratory values (white blood cell count, hemoglobin, platelet count, glucose levels), comorbidities (cardiovascular disease, hypertension), and treatment-related factors (length of stay, ventilation usage). Clinical experts in ICU care then reviewed and validated these selected features, confirming their practical relevance and clinical significance. The complete feature list is illustrated in Table1.

**Table 1.**
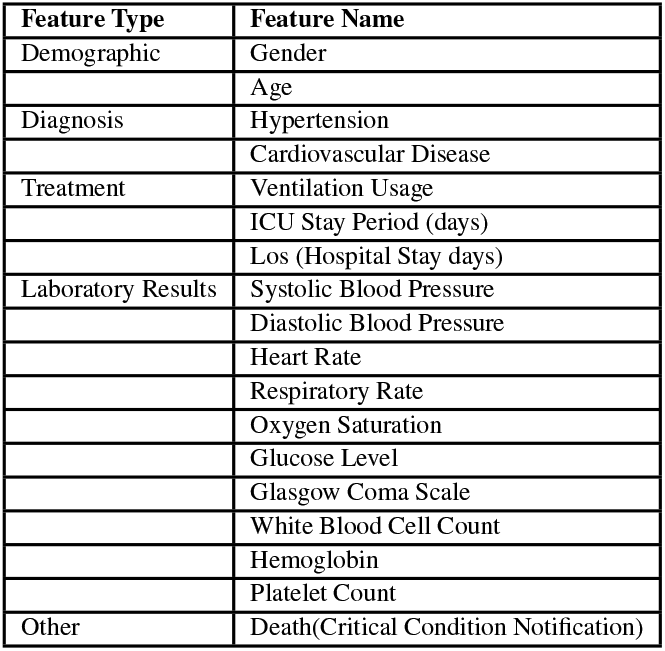
Feature List.

### 2.4. Preprocessing Method

Our data preprocessing pipeline consisted of several key steps to ensure data quality and model performance. First, we addressed data completeness by removing columns with over 90% missing values, while applying mean imputation for the remaining missing values.

For categorical variables, we employed Label Encoder to transform them into numerical format, enabling their integration into our machine learning models. This transformation was essential for features such as gender, race, and various clinical conditions.

Given the inherent class imbalance in ICU readmission data, we implemented the RandomUnderSampler technique to balance our dataset [24]. This approach reduced the number of majority class samples to 60% of the minority class, helping to prevent bias in model training while maintaining data integrity.

### 2.5. Modeling

In our study of 2,144 patients with 18 features, we employed a 70/30 train-test split for model evaluation. Multiple machine learning algorithms were implemented, including Random Forest, XGBoost, Light GBM, and AdaBoost, to develop predictive models.

To facilitate comprehensive model comparison, we evaluated four different machine learning algorithms. Random Forest [27], which combines multiple decision trees through bootstrap aggregation for robust predictions, was included alongside XGBoost and LightGBM - both gradient boosting [28] frameworks that sequentially build trees to minimize prediction errors, with LightGBM specifically employing a leaf-wise growth strategy [29]. AdaBoost, as an ensemble learning method, combines multiple weak learners sequentially, with each subsequent model focusing on previously misclassified samples.

We optimized these models using a specific parameter space tailored to each model. We defined common hyperparameters such as the number of estimators, the learning rate, and maximum depth, with LightGBM additionally including the number of leaves parameter. We implemented an optimization function using the Optuna library for hyperparameter tuning. The function contains an optimization objective that uses different parameters within its defined range. The goal of Optuna study was to maximize the AUROC scores through multiple trials, thoroughly exploring the parameter space. We applied RFECV to select the most important features. The model, with the suggested parameters, is fitted on the training data. RFECV helps identify and select the most relevant features tailored to the specific model, which are then used to train the model.

Model performance was evaluated using multiple metrics, with AUROC selected as the primary evaluation criterion due to its distinct advantages. AUROC effectively handles class imbalance and provides a threshold-independent assessment of model discrimination. Additionally, its widespread use in medical prediction literature facilitates meaningful comparisons with existing studies [25]. Complementary metrics including accuracy, sensitivity, and specificity were also calculated to ensure comprehensive performance assessment.

Through systematic comparison, AdaBoost emerged as our proposed model due to its superior performance. For this model, we achieved optimal performance with a learning rate of 0.178 and 253 estimators. The base estimator was configured as a decision tree with maximum depth of 1, striking a balance between model complexity and generalization capability. These parameters were determined through extensive cross-validation and optimization trials, with random state set to 123 to ensure reproducibility. The RFECV implementation utilized 5-fold cross-validation with the same AdaBoost configuration as the estimator.

AdaBoost’s effectiveness can be attributed to several key characteristics. Its key advantages include robust handling of outliers, effective prevention of overfitting through iterative learning, and strong generalization capabilities [26]. The algorithm’s adaptive weighting mechanism allows it to give more importance to difficult-to-classify cases, which is especially valuable in medical prediction scenarios where identifying high-risk cases is crucial. Its iterative learning process, which focuses on previously misclassified samples, makes it particularly adept at handling complex medical data patterns.

### 2.6. Statistical Analysis of Models

To validate the statistical robustness of our model results, we conducted comprehensive statistical analyses comparing the train and test sets using both t-tests and chi-square tests. The dataset was split into a 70% training set and a 30% test set for model evaluation. T-tests were employed to compare the means of continuous variables between the train and test sets, while chi-square tests were utilized to assess the independence of categorical variables between the two sets. A threshold p-value of 0.05 was established to determine statistical significance [32]. We tested the hypothesis that there was no significant difference between the train and test sets. The results showing p-values greater than 0.05 indicated no statistically significant differences between the datasets, confirming the consistency and generalizability of our model’s performance.

### 2.7. Feature Importance

We employed SHAP (SHapley Additive exPlanations) values to understand how each feature influences our model’s predictions. SHAP values are particularly useful in medical applications as they provide clear and interpretable insights into model behavior, helping healthcare professionals understand the reasoning behind predictions [33, 34, 35].

Additionally, we conducted a feature ablation study, which systematically evaluates the model’s performance by removing one feature at a time. This technique helps understand each feature’s contribution by measuring the decrease in AUC score when that feature is excluded - a larger decrease indicates greater feature importance [36].

## 3. RESULTS

### 3.1. Cohort Comparison

Following our approach for feature selection and data preprocessing discussed earlier, our final dataset included 2,144 patients from MIMIC-III and MIMIC-IV databases. The selected cohort was randomly split into train/test sets with a ratio of 70/30, resulting in 1,500 patients for training and 644 for testing.

The demographic analysis revealed comparable distributions between cohorts across a variety of features. The median age was 69.0 years (IQR: 15.2) in the training cohort and 69.0 years (IQR: 16.1) in the testing cohort, with no statistically significant difference between them (p = 0.89). Gender distribution showed similar patterns, with males comprising 56% (835 patients) of the training cohort and 55% (352 patients) of the testing cohort (p = 0.45). Clinical characteristics included a range of vital signs and laboratory results, which were consistent across cohorts. Systolic blood pressure averaged 152±39.5 mmHg in the training cohort and 154±38.4 mmHg in the testing cohort (p = 0.27), while diastolic blood pressure was 99±19.1 mmHg and 99±9.7 mmHg, respectively (p = 0.31). Heart rate and respiratory rate also showed no significant differences, with values of 97±27.6 bpm and 25±12.5 in the training cohort, and 96±27.3 bpm and 25±11.3 in the testing cohort (p = 0.95 and p = 0.33, respectively). Oxygen saturation levels were similar across groups (99.0±1.3 in training and 99±1.6 in testing, p = 0.40), as were glucose levels (136±71.9 mg/dL in training, 136±81.3 mg/dL in testing, p = 0.32). Additional laboratory measures, including hemoglobin (13±2.3 g/dL in both cohorts, p = 0.16), platelet count (288±133.0 K/uL in training, 288±136.6 K/uL in testing, p = 0.13), and white blood cell count (9.8±5.4 and 9.8±13.0, p = 0.32) showed no statistical differences. The Glasgow Coma Scale (GCS) scores were also comparable, with an average of 15±1.8 in the training cohort and 15±2.0 in the testing cohort (p = 0.40). Regarding comorbidities, both cohorts had similar rates of hypertension (44% in training vs. 43% in testing, p = 0.79) and cardiovascular disease (37% vs. 38%, p = 1). The ICU stay period averaged 3 days (±3.43 in training and ±3.17 in testing, p = 0.63), the hospital stay period averaged 3 days(±7.3 in training and ±7.4 in testing, p = 0.67). This suggests that some patients may have been transferred to the ICU later during their hospital stay. Ventilation usage rates were 34% in the training cohort and 33% in the testing cohort (p = 0.88). Mortality rates were also consistent, with 8% (125 patients) in the training cohort and 11% (70 patients) in the testing cohort (p = 0.94). Finally, ICU readmission status was similar across cohorts, with 3% in the training and testing cohorts (p = 1.00). Statistical analyses using t-tests for continuous variables and chi-square tests for categorical variables (p > 0.05 threshold) confirmed the comparability of most features between cohorts. The comprehensive statistical assessment, as detailed in Table2, validated the robustness of our data splitting strategy and supported the generalizability of our model’s results.

**Table 2.**
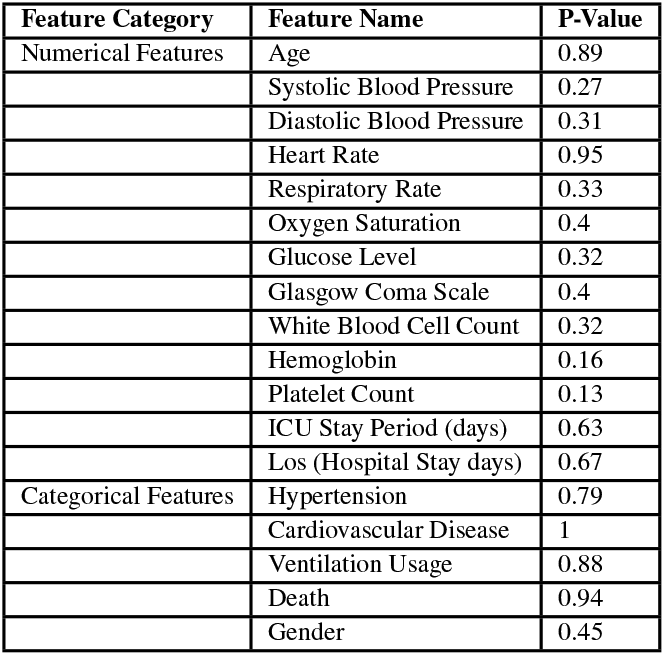
Cohort Comparison.

| Feature Category | Feature Name | P-Value |
| --- | --- | --- |
| Numerical Features | Age | 0.89 |
|  | Systolic Blood Pressure | 0.27 |
|  | Diastolic Blood Pressure | 0.31 |
|  | Heart Rate | 0.95 |
|  | Respiratory Rate | 0.33 |
|  | Oxygen Saturation | 0.4 |
|  | Glucose Level | 0.32 |
|  | Glasgow Coma Scale | 0.4 |
|  | White Blood Cell Count | 0.32 |
|  | Hemoglobin | 0.16 |
|  | Platelet Count | 0.13 |
|  | ICU Stay Period (days) | 0.63 |
|  | Los (Hospital Stay days) | 0.67 |
| Categorical Features | Hypertension | 0.79 |
|  | Cardiovascular Disease | 1 |
|  | Ventilation Usage | 0.88 |
|  | Death | 0.94 |
|  | Gender | 0.45 |

### 3.2. Model Assesment

The summary of the results for both proposed and baseline models are shown in Table3. The AdaBoost model emerged as the champion, demonstrating superior performance with an AUC of 0.877 (95% CI = [0.815-0.913]), accuracy of 0.810, sensitivity of 0.843, and specificity of 0.802. The performance metrics underscore not only its accuracy but also its robustness in balancing between Type I and Type II errors, particularly in identifying high-risk patients requiring readmission. In addition to these primary metrics, we conducted a comprehensive evaluation of model performance across all tested algorithms. Random Forest and XGBoost achieved comparable performance with AUROC values of 0.841 and 0.865 respectively, while LightGBM showed slightly lower performance with an AUROC of 0.828. The ROC curve analysis, as illustrated in Fig2, provides a visual representation of these results, highlighting the superior performance of AdaBoost across different threshold settings.

**Table 3.** Result Metrics for Proposed and Baseline Models.

| Model | Accuracy | Sensitivity | Specificity | AUROC | 95% CI |  |
| --- | --- | --- | --- | --- | --- | --- |
| Adaboost | 0.810 | 0.843 | 0.802 | 0.877 | 0.815 | 0.913 |
| XGBoost | 0.771 | 0.840 | 0.761 | 0.865 | 0.812 | 0.905 |
| Random Forest | 0.782 | 0.840 | 0.782 | 0.841 | 0.801 | 0.886 |
| Light GBM | 0.714 | 0.792 | 0.710 | 0.828 | 0.775 | 0.885 |

**Figure 2.**
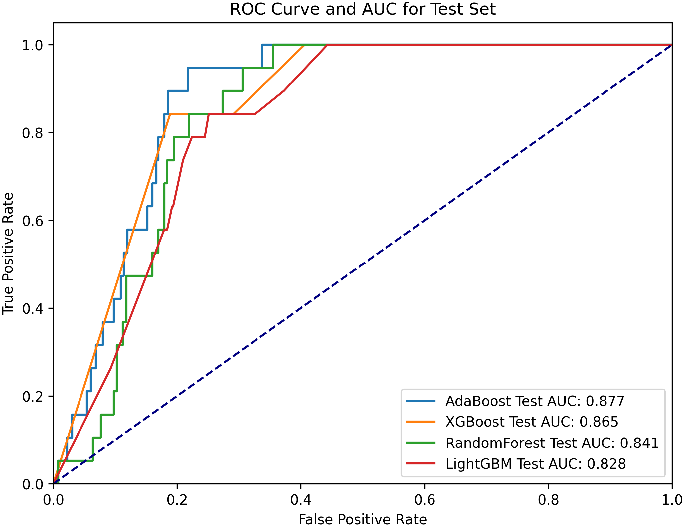
AUROC Curve for Proposed and Baseline Models

The AdaBoost model’s ensemble approach, combining multiple weak learners sequentially, provides robust generalization capabilities while maintaining computational efficiency [30, 31]. Each subsequent model in the ensemble focuses on previously misclassified samples, making it particularly effective in handling complex medical data patterns. This adaptive weighting mechanism allows it to give more importance to difficult-to-classify cases, which is especially valuable in medical prediction scenarios where identifying high-risk cases is crucial [26]. Furthermore, AdaBoost consistently outperformed other models across all evaluation criteria, demonstrating stable performance in both majority and minority classes. This balanced performance is particularly important in clinical settings, where both false positives and false negatives can have significant implications for patient care and resource allocation.

### 3.3. SHAP analysis

The SHAP value analysis, illustrated in Fig**??** provides comprehensive insights into feature importance and their relationships with ICU readmission predictions. Our analysis reveals that white blood cell count emerged as the most statistically significant predictor, followed by cardiovascular disease and hemoglobin. The relationship between white blood cell count and readmission risk is notably strong, as evidenced by the longest bars in both Class 0 and Class 1 in the bar plot, and the wide distribution of SHAP values in the summary plot. Cardiovascular disease shows substantial impact on readmission predictions, ranking second in importance. The SHAP values indicate that the presence of cardiovascular disease is strongly associated with increased readmission risk, as shown by the concentration of red dots on the positive SHAP value side. Hemoglobin and GCS (Glasgow Coma Scale) demonstrated similar levels of predictive power, ranking third and fourth in importance. The summary plot reveals that extreme values in these parameters contribute significantly to readmission predictions. Platelet count and gender also emerge as important predictors, though with moderate impact compared to the top features. These hematologic parameters and demographic factors play a supportive but meaningful role in predicting readmission, suggesting that the model captures both physiological and patient-specific characteristics. The analysis also highlights the contributions of length of stay (los) and age, which provide important contextual information about patient status and vulnerability. Interestingly, clinical parameters traditionally considered crucial, such as blood pressure (both systolic and diastolic), heart rate, and oxygen saturation, showed relatively lower SHAP values in our analysis. This suggests that while these vital signs remain clinically relevant, they may serve more as supporting indicators rather than primary drivers of readmission risk in our model context. By leveraging these SHAP insights, we identified that hematological parameters (white blood cell count, hemoglobin, platelet count) and chronic conditions (cardiovascular disease) are the strongest predictors of ICU readmission. This finding suggests that underlying systemic conditions and chronic diseases might be more predictive of readmission risk than acute physiological measurements, potentially offering new perspectives for clinical risk assessment and intervention strategies.

### 3.4. Ablation Study on Variable

Feature ablation analysis, as illustrated in Fig4, revealed that gcs and cardiovascular disease are fundamental to our model’s predictive capability, with their removal causing the most substantial degradation in model performance (AUROC dropping to approximately 0.81-0.82).

**Figure 3.**
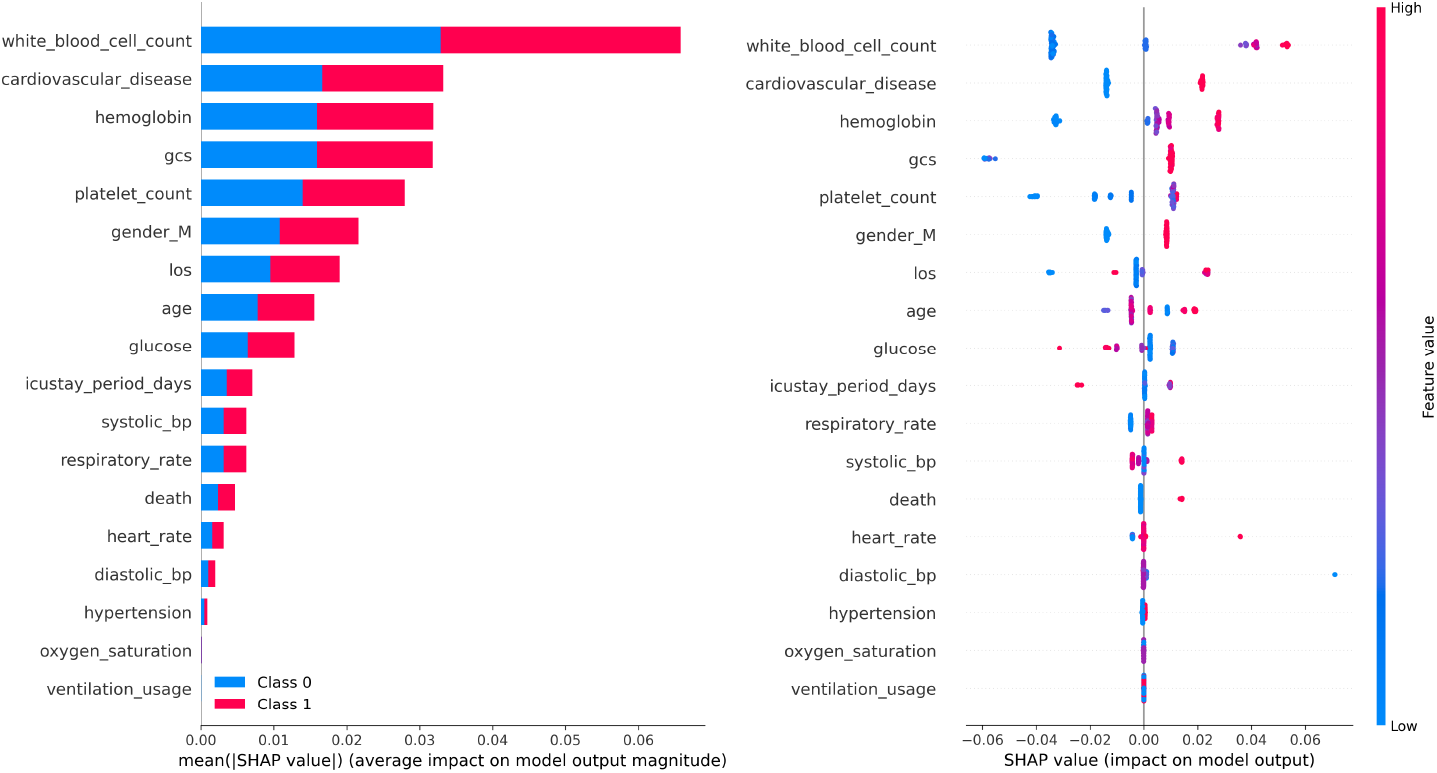
SHAP Bar Analysis for Adaboost Model

**Figure 4.**
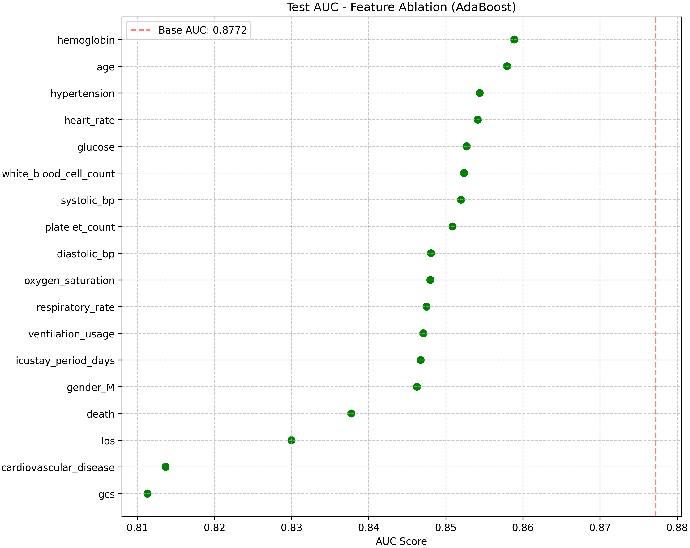
Ablation Study for Adaboost Proposed Model

This finding suggests these features contain unique predictive information that cannot be fully compensated by other variables. Most other variables, including hemoglobin, age, hypertension, and heartrate, showed relatively minor impact on model performance when removed individually, maintaining AUROC scores around 0.85-0.86. Interestingly, clinical parameters such as respiratory rate, oxygen saturation, and ventilation usage demonstrated moderate importance to the model’s performance. This systematic evaluation of feature importance through ablation testing helps validate our feature selection process and suggests that while certain acute physiological parameters (like gcs) are crucial for prediction, the model relies on a balanced combination of both acute measurements and chronic conditions (such as cardiovascular disease) in assessing ICU readmission risk.

## 4. DISCUSSION

### 4.1. Existing model compilation summary

Several methods have been developed to predict ICU readmission for ICH patients using MIMIC databases. The most relevant previous study represented in Table4 achieved an AUC of 0.736 (95% CI: 0.668-0.801) and an accuracy of 86.2% using LightGBM model [12].

**Table 4.** Comparative Analysis between Our Study and the Best Existing Study.

| Proposed Model | LightGBM | AdaBoost |
| --- | --- | --- |
| AUC | 0.736 | 0.877 |
| 95% CI for AUC | [0.668, 0.801] | [0.815, 0.913] |
| Feature Engineering Techniques | SMOTEEN, Bayesian Optimization, RFECV | RandomUnderSampler, Optuna framework, RFECV |
| Top 5 Significant Features | Hydrocephalus, Sex, Neutrophils, Glasgow Coma Scale (GCS), Specific Oxygen Saturation (SpO2) levels | White Blood Cell Count, Cardiovascular Disease, Hemoglobin, Glasgow Coma Scale (GCS), Platelet Count |

In our study, we implemented several innovative approaches, particularly focusing on the challenge of class imbalance using the RandomUnderSampler technique. Our feature selection process, combining RFECV with clinical expertise, helped identify 18 key features from an initial set of 50 potential predictors. The AdaBoost model, with its adaptive weighting mechanism, proved especially effective in handling imbalanced datasets and focusing on misclassified samples.

Although the existing methodology was successful in achieving high accuracy, it possessed several limitations. First, they failed to effectively address the class imbalance issue, resulting in low sensitivity scores around 22.6%, indicating poor performance in identifying patients who actually needed ICU readmission. Second, while they conducted SHAP analysis, their approach lacked complementary feature importance validation methods, potentially limiting the robustness of their feature importance findings. Additionally, their model showed limited ability to identify high-risk patients requiring readmission.

Our proposed approach offers several advantages over prior research:

- Novel preprocessing techniques using RandomUnderSampler significantly enhanced model performance, achieving substantial improvements in identifying patients requiring readmission (sensitivity from 22.6% to 84%) while maintaining a high AUC of 0.877
- Enhanced feature importance analysis by complementing SHAP with ablation studies, providing a more comprehensive and validated understanding of feature impacts
- Our approach to addressing class imbalance led to more clinically relevant predictions, particularly enhancing the identification of high-risk patients requiring readmission
- Utilization of AdaBoost and advanced feature selection processes (RFECV) provided a more robust framework for handling imbalanced medical data with better interpretability

These advancements, compared to [12], represent significant improvements in ICU readmission prediction for ICH patients. While trading off some overall accuracy (81% vs their 86.2%), our model achieved substantially better AUC (0.87 vs 0.736) and sensitivity (84% vs 22.6%), demonstrating superior ability in identifying patients who truly need ICU readmission and resulting in a more balanced and clinically applicable approach.

### 4.2. Limitations and Future Work

Firstly, since both MIMIC-III and MIMIC-IV are single-center databases, the generalizability of our AdaBoost model across different clinical settings may be limited. Furthermore, the reliance on ICD-9 and ICD-10 codes to identify patients with intracerebral hemorrhage (ICH) introduces variability, as these codes represent broad classifications. Patients may have different underlying causes of ICH, such as trauma or spontaneous hemorrhage, which we were unable to analyze in detail. To enhance the model’s generalizability, future research should explore validating the performance of the AdaBoost model using multi-center databases and further investigate the relationship between different causes of ICH and ICU readmissions to improve the model’s predictive accuracy and clinical applicability.

Secondly, although this study employed advanced techniques for feature selection and data preprocessing, there is still room for improvement in handling missing data and class imbalance within the AdaBoost model. Future research could explore more sophisticated imputation methods, such as multiple imputation or other machine learning-based techniques, to minimize the impact of missing data on model performance. Additionally, while AdaBoost’s ability to automatically adjust sample weights provides a certain advantage in addressing class imbalance, future work could integrate different balancing techniques to further enhance the classification performance for minority class samples.

Additionally, while AdaBoost performed well in this study, future research could benefit from integrating deep learning approaches, such as Convolutional Neural Networks (CNNs) and Recurrent Neural Networks (RNNs), to handle more complex and high-dimensional data. Moreover, this study utilized average values of laboratory data as indicators of patient condition, which overlooked important temporal information. Incorporating time-series analysis in future work could capture data fluctuations and trends, thereby further improving the predictive power and generalizability of the AdaBoost model.

Furthermore, the implementation of predictive models in clinical practice is of critical importance. Developing user-friendly tools and interfaces that can be seamlessly integrated into clinical workflows is key to applying the predictive outcomes of the AdaBoost model in practice. Collaborating with clinicians during the development process to understand their needs and preferences will ensure that the AdaBoost model is not only accurate but also easy to operate in practical settings. Additionally, understanding privacy and ethical concerns when handling sensitive health data is crucial to fostering trust in the model’s use in clinical practice.

Lastly, future research should also explore the possibility of utilizing real-time data streams and continuous learning models to ensure that the AdaBoost model remains updated with the latest clinical data. This dynamic model adjustment approach would improve the responsiveness and accuracy of the AdaBoost model, ensuring its continued relevance and effectiveness in rapidly evolving clinical environments, particularly in complex ICU settings.

## 5. CONCLUSION

This study successfully developed and validated a machine learning framework to predict ICU readmissions in intracerebral hemorrhage patients through the MIMIC databases. Our comprehensive approach, combining RandomUnderSampler technique, advanced feature selection through RFECV, and optimization of the AdaBoost model, has demonstrated significant improvements in predictive performance. Most notably, we achieved a remarkable improvement in sensitivity from 22.6% to 84.3%, while maintaining a strong AUROC of 0.877 (95% CI: 0.815-0.913). This advancement in identifying high-risk patients, despite a slight trade-off in overall accuracy, represents a significant step forward in clinical applicability. Through the integration of dual feature importance analysis methods - SHAP and ablation studies - our approach not only enhanced model interpretability but also provided robust validation of feature selection. These methodological innovations have improved both computational efficiency and model generalization to unseen data, demonstrating the potential of machine learning as a valuable tool in dynamic ICU environments where precise predictive models are crucial. The immediate clinical applicability of our framework lies in its ability to assist healthcare providers in making informed decisions about ICU discharge timing and post-discharge monitoring intensity. Our model’s high sensitivity in identifying high-risk patients can help prevent premature discharges and optimize resource allocation. Healthcare institutions can implement this framework alongside existing clinical protocols to provide data-driven support for decision-making, particularly in cases where readmission risk assessment is challenging. Future research directions should focus on validating our approach using datasets from diverse healthcare systems and exploring additional data types available in MIMIC databases, such as clinical notes and temporal patterns. The integration of these varied data sources could provide richer context and potentially lead to even more accurate and reliable predictions. Furthermore, real-time data integration and multi-center validation would be essential steps toward developing implementable clinical decision support tools. Ultimately, through these continued research efforts and methodology refinements, we anticipate further enhancing the model’s generalizability and clinical utility. This ongoing development is crucial for creating reliable tools that can effectively support clinical decision-making, thereby improving patient outcomes in intensive care settings.

## Declarations

## Acknowledgment

The authors extend their gratitude to the creators of MIMIC-III and MIMIC-IV for furnishing a thorough and inclusive public electronic health record (EHR) dataset.

During the preparation of this work the H.L. used CLAUD in order to improve the readability and language of the manuscript. After using this tool, the H.L, M.P reviewed and edited the content as needed and take full responsibility for the content of the published article.

Special thanks to Negin Ashrafi for her constructive feedback and suggestions that contributed to the improvement of this manuscript.

## Author Contributions

H.L., R.M., M.P. : Involved in all aspects of this study. E.P.:Provided expert insights.

## Data Availability

The Medical Information Mart for Intensive Care III (MIMIC-III) is a comprehensive dataset, available to the public via https://physionet.org/content/mimiciii/1.4/.

The Medical Information Mart for Intensive Care IV (MIMIC-IV) is a comprehensive dataset, available to the public via https://physionet.org/content/mimiciv/2.0/

## Ethics Approval and Consent to Participate

The dataset used to support the conclusions of this article is sourced from the Medical Information Mart for Intensive Care version III and IV(MIMIC-III, MIMIC-IV). As these databases are public and de-identified, informed consent and Institutional Review Board approval were not required. All procedures followed the relevant guidelines and regulations.

## Consent for Publication

Not applicable.

